# Monitoring tubular epithelial cell damage in AKI via urine flow cytometry

**DOI:** 10.1101/2022.01.31.22270101

**Authors:** Jacob Kujat, Valerie Langhans, Hannah Brand, Paul Freund, Nina Görlich, Leonie Wagner, Diana Metzke, Sara Timm, Matthias Ochs, Andreas Grützkau, Sabine Baumgart, Christopher M. Skopnik, Falk Hiepe, Gabriela Riemekasten, Jan Klocke, Philipp Enghard

**Affiliations:** Department of Nephrology and Medical Intensive Care, Charité-Universitätsmedizin Berlin, corporate member of Freie Universität Berlin and Humboldt-Universität zu Berlin, Berlin, Germany; German Rheumatism Research Center Berlin (DRFZ), an Institute of the Leibniz Foundation, Berlin, Germany; Core Facility Electron Microscopy, Charité-Universitätsmedizin Berlin, corporate member of Freie Universität Berlin and Humboldt-Universität zu Berlin, Berlin, Germany; Institute of Functional Anatomy, Charité-Universitätsmedizin Berlin, corporate member of Freie Universität Berlin and Humboldt-Universität zu Berlin, Berlin, Germany; German Center for Lung Research (DZL), Berlin, Germany; Institute for Immunology/Core Facility Cytometry, Jena University Hospital, Jena, Germany; Department of Rheumatology and Clinical Immunology, Charité-Universitätsmedizin Berlin, Berlin, Germany; Department of Rheumatology, University of Lübeck, Lübeck, Germany

## Abstract

Acute kidney injury (AKI) is associated with significant morbidity and mortality. The diagnosis is currently based on urine output and serum creatinine and there is a lack of biomarkers that directly reflect tubular damage. Here, we establish flow cytometric quantification of renal epithelial cells as a potential biomarker for quantifying the severity of tubular kidney damage and for predicting AKI outcome.

**Methods:** A total of 84 patients with AKI were included in this study, divided into an exploratory cohort (n=21) and confirmatory cohort (n=63), as well as 25 controls. Urine of patients was collected and processed within 72 hours after AKI onset. Different urinary tubular epithelial cell (TEC) populations were identified and quantified by flow cytometry (FACS). Urinary cell counts were analyzed regarding AKI severity defined by KDIGO stage as well as renal recovery, length of hospital stay and occurrence of MAKE-30 events.

**Results:** Urinary TEC counts correlated with stages of AKI based on KDIGO classification and were significantly enriched in patients with AKI compared to healthy donors and inpatient controls in both cohorts. Furthermore, both proximal and distal TEC (pTEC, dTEC) counts performed well in identification of patients with AKI regardless of stage. Urinary amounts of pTEC and dTEC showed a strong correlation, with predominance of dTEC. Higher numbers of TEC were associated with extended length of hospital stay, while elevated pTEC counts were associated with the occurrence of MAKE-30 events. Follow-up measurements showed decreasing amounts of urinary TEC after AKI recovery over several days.

**Conclusion:** The amount of urinary TEC directly reflects severity of tissue damage in human AKI. Our protocol furthermore provides a basis for a deeper phenotypic analysis of urinary TEC populations.

## Introduction

Acute kidney injury (AKI) is characterized by an acute impairment of kidney function associated with a high morbidity and hospitalization rate.^1^ Affecting up to 20% of hospitalized patients, AKI is a common problem in clinical settings and is associated with increased mortality.^2^ Furthermore, even after recovery, patients with AKI have an increased risk of developing subsequent chronic kidney disease (CKD).^3^

Currently the diagnosis and stratification of AKI is based on changes in serum creatinine (SCr) and/or urine output.^4–6^ However, SCr is a marker for kidney function and changes only take place when glomerular function has decreased by approximately 50%. Therefore, it stands at the very end of a chain of complex damaging mechanisms.^7^ Novel biomarkers reflecting disturbances upstream of functional impairment may facilitate earlier diagnosis of AKI and more direct assessment of kidney damage.

Several molecules have been investigated as potential biomarkers for early diagnosis in AKI patients. Promising results have been reported for different markers of kidney failure such as Cystatin C and markers of structural kidney damage such as IL-18,^8^ CCL-14,^9^ Neutrophil Gelatinase-Associated Lipocalin (NGAL)^8,10^ and Kidney Injury Molecule 1 (KIM-1)^11^. Moreover, the combination of cell cycle arrest biomarkers TIMP-2 and IGFBP-7 (Nephrocheck®) has been validated to identify critically ill patients that are prone to AKI development.^12,13^ However, novel structural damage markers (CCL-14, KIM-1, Nephrocheck®, etc.) have not yet made the translational leap to the patient’s bedside.

Detection of tubular epithelial cells (TEC) and cell casts via urine microscopy is a well-established marker for acute tubular necrosis (ATN), a characteristic event in most forms of AKI.^14,15^ Moreover, high amounts of renal TEC or cell casts in urine have been associated with need for dialysis,^16^ probability of nonrecovery,^17^ adjusted odds of worsening AKI and sepsis.^18^ The use of microscopy however is impaired by low interobserver reliability, lack of clinician experience and time constraints in modern clinical practice. Nonetheless, quantifying urinary TEC as the pathophysiological correlate of structural kidney damage on a cellular level remains a reasonable approach for a biomarker of structural kidney injury.

FACS provides an observer independent approach for detection and quantification of urinary TEC. Our group previously reported the usage of urinary T cells (and TEC) as a powerful biomarker for intrarenal inflammation in lupus nephritis and kidney transplant rejection.^19,20^ We assumed that the amount of urinary TEC measured via FACS is a suitable biomarker for the severity of kidney damage. We hypothesized that the amount of urinary TEC increases in patients with more severe AKI, as severe tubular injury leads to detachment of TEC from their basement membrane^21^. Furthermore, we speculated whether separately quantifying proximal und distal TEC may yield additional information on the severity of AKI. Finally, we assessed the ability of urinary TEC to predict AKI outcome parameters such as duration of AKI until recovery, timespan of hospital stay and major adverse kidney events within 30 days (MAKE-30).^22^

## Methods

For this trial, a proof-of-concept exploratory study and a subsequent confirmatory study were performed on a cross-sectional cohort of patients with AKI. Urine samples were collected from June 2015 to December 2016 (exploratory study) and from April 2018 to August 2019 (confirmatory study). All samples were donated by patients of the Department of Nephrology and Intensive Care Medicine, Cardiology or Surgery, Charité University Hospital, Berlin.

AKI was defined by applying the KDIGO criteria^5^, which requires an increase of SCr by a factor of at least 1.5 within 7 days or an increase by at least 0.3 mg/dL within 48 hours to classify a kidney injury as AKI. If SCr values of the 7 days prior to AKI were not available, the lowest SCr value of the 365 days after recovery from AKI or, preferably, the lowest SCr value of the 365 days before AKI onset was used as baseline surrogate. In two cases of the confirmatory study, baseline values calculated from an eGFR of 75ml/min/1.73m^2^ had to be used.

### Patients and routine laboratory values

We collected and analyzed urine from 84 patients with AKI and 25 controls, divided into an initial exploratory cohort (21 AKI patients) and a subsequent confirmatory cohort (63 AKI patients). The inclusion criterion in both studies was the presence of AKI with clear trigger (sepsis, cardial decompensation, volume depletion) without suspicion of glomerulonephritis or acute interstitial nephritis within 72 hours before urine collection. Patients with urinary tract infections or postrenal AKI were excluded from analysis, as well as children and kidney transplanted patients.

All samples were collected within 72 hours after onset of kidney damage and examined using FACS. All patients were classified according to their maximum KDIGO stage within 72 hours after AKI onset (referred to as KDIGO groups “1”, “2”, or “3”). Patient characteristics were collected using pre-existing data from the electronic health software SAP and are summarized in *table 1*. Since the available data did not include urine output, only SCr values were used for AKI classification. All SCr values, including follow-up data, were retrieved from the electronic medical records as well. SCr values were measured using the Jaffé method. Renal recovery was defined according to the definition of AKI as renormalization of the SCr below a value 1.5 times the baseline value or below the baseline value plus 0.3 mg/dL, depending on the AKI subdefinition used for study subject inclusion. Occurrence of MAKE-30 event was defined as persistently impaired kidney function, new renal replacement therapy, or in-hospital mortality within 30 days following AKI onset. Duration was calculated using the date of AKI onset and the date of the first SCr value within the recovery range as stated above (if recovery occurred), the date of exitus (if the patient died) or the last date of SCr analysis documented in the electronic health software system (if recovery did not occur).

**Table 1.** Characteristics of patients. Values are mean (range). Data for AKI are from 21 patients in the exploratory study and 63 patients in the confirmatory study. CKD, chronic kidney disease; HC, healthy controls; IPC, inpatient controls; KDIGO, Kidney Disease: Improving Global Outcomes, AKI groups; Np, not performed; Na, not applicable; SCr, serum creatinine.

|  | Exploratory study |  |  |  |  | Confirmatory study |  |  |  |  |
| --- | --- | --- | --- | --- | --- | --- | --- | --- | --- | --- |
|  | Controls |  | KDIGO |  |  | Controls |  | KDIGO |  |  |
|  | HC | IPC | 1 | 2 | 3 | HC | IPC | 1 | 2 | 3 |
| *n* | 5 | 5 | 7 | 5 | 9 | 5 | 10 | 35 | 13 | 15 |
| *Age (years)* | 35 (23-51) | 63 (41-74) | 67 (64-77) | 65 (52-80) | 61(40-82) | 37 (24-51) | 69 (56-89) | 70 (40-86) | 69 (34-84) | 66 (36-80) |
| *Female/Male* | 2/3 | 2/3 | 2/5 | 1/4 | 4/5 | 1/4 | 7/3 | 9/26 | 3/10 | 8/7 |
| *SCr on day of measurement (mg/dl)* | Np | 0,48 (0.25-0.76) | 1,29 (0.69-1.69) | 1,96 (1.3-2.8) | 2,22 (0.93-4.15) | Np | Np | 1.40 (0.98-1.77) | 1.88 (1.22-3.39) | 2.28 (0.97-4.23) |
| *Etiology of prerenal AKI*<br>*Cardiogenic/Septic/Other)* | Na | Na | 4/3/0 | 3/1/1 | 5/4/0 | Na | Na | 18/2/15 | 5/4/4 | 4/6/5 |
| *Pre-existing CKD (yes/no)* | 0/5 | 0/5 | 2/5 | 1/4 | 1/8 | 0/5 | 0/5 | 10/25 | 2/11 | 1/14 |
| *MAKE-30 (yes/no)* | 0/5 | 0/5 | 4/3 | 4/1 | 7/2 | 0/5 | 0/10 | 6/29 | 3/10 | 7/8 |

The control groups for both studies consisted of 5 healthy individuals without previous renal impairment (referred to as “healthy controls” or “HC”) and 5 (exploratory cohort) or 10 (confirmatory cohort) inpatients without current renal impairment (referred to as “inpatient controls” or “IPC”).

Written informed consent was obtained by all patients before study participation. Renal and ureteral tissue samples were derived from deceased patients, written consent was obtained by the patient’s relatives. Ethical approval was given by the ethics committee of Charité University Hospital (Charité EA1/239/16 and EA2/045/18). The studies were carried out according to the ethical guidelines of the Helsinki Declaration and our institution.

### Establishing a staining protocol for the detection of tubular epithelial cells

We established a staining protocol for urinary pTEC and dTEC using human post-mortem renal and ureteral tissue samples. Four kidney samples and one ureter sample were stored in Gibco Roswell Park Memorial Institute (RPMI) 1640 Medium (Thermo Fisher Scientific, Waltham, USA) containing 10% Fetal Calf Serum (FCS) and 1% Penicillin/Streptomycin (P/S) and digested using Collagenase VIII (1,5 mg/ml, Serva Electrophoresis GmbH, Heidelberg, Germany) and DNAse I (50µg/ml, Roche, Basel, Switzerland). The staining protocol and the utilized antibodies correspond with the urinary staining protocol for confirmatory cohort as expounded below.

### Sample preparation and flow cytometry

After collection, urine samples were immediately centrifuged and washed in phosphate-buffered saline containing 0,5% bovine serum albumin (PBS-BSA) and 2mM ethylenediaminetetraacetic acid (EDTA; Thermo Fisher Scientific, Waltham, USA). Approximate standing time for the urine before preparation in both studies was 3-6 hours. First-morning void urine samples were used preferentially. For staining with cytokeratin, cells were fixed using 4% paraformaldehyde and subsequently perforated using Saponin (exploratory cohort; Sigma-Aldrich, Hamburg, Germany, 5mg/mL in PBS) or Perm/Wash solution (confirmatory cohort; BD Biosciences, Franklin Lakes, USA). Cells were then resuspended in PBS/BSA containing 10% human IgG (Flebogamma; Grifols, Langen, Germany) to block unspecific binding. Afterwards, staining with antibodies listed below was carried out by incubation for 20 minutes at 4°C in the dark. Samples were filtered immediately before analysis using MACS pre-separation filters with a mesh size of 70µm. If cell sorting prior to microscopic examination was performed, a mesh size of 30µm was used (Miltenyi Biotec GmbH, Bergisch Gladbach, Germany). In the confirmatory study, 4ʹ,6-Diamidine-2ʹ-phenylindole dihydrochloride (DAPI; Sigma-Aldrich, Hamburg, Germany) was added immediately before FACS analysis to exclude non-DNA-containing urinary debris and thereby reduce background noise, as this is a major problem in FACS analyses of human urine. Stained samples were processed using a MACS Quant Analyzer (exploratory cohort; Miltenyi Biotec GmbH, Bergisch Gladbach, Germany) or a BD FACSCanto II Flow Cytometer, respectively (confirmatory cohort; BD Biosciences, Franklin Lakes, USA). Final analysis was carried out using FlowJo Software (Tree Star, Ashland, Oregon, USA). Calculated cell numbers were normalized to a volume of 100 mL urine.

### Antibodies

We employed a combination of markers based on literature research for the distinct detection of proximal TEC as well as distal TEC. CD10 (= neutral endopeptidase or NEP) and CD13 (= alanine aminopeptidase or AAP)^23,24^ were used to identify proximal TEC, CD227 (= mucin 1 or MUC-1)^25^ and CD326 (= epithelial cell adhesion molecule or EpCam)^26^ to detect distal TEC and both marker combinations included a pan-cytokeratin marker.^27^ The staining protocol of the exploratory study included CD10-PE-Vio770 (Clone: 97C5, Isotype: Mouse IgG1, Miltenyi Biotec GmbH, Bergisch Gladbach, Germany), CD326-PE (Clone: HEA-125, Isotype: mouse IgG1k, Miltenyi Biotec GmbH, Bergisch Gladbach, Germany and pan-cytokeratin-Alexa Fluor 647 (Clone: C-11, Isotype: mouse IgG1k, BioLegend, San Diego, USA). After proving the feasibility of this method in the exploratory study, the established marker combination was improved by adding CD13 and CD227 to the existing staining panel. The complete staining protocol included pan-cytokeratin-APC (Clone: REA831, Isotype: Recombinant human IgG1) or, exclusively for fluorescence microscopy, pan-cytokeratin-FITC (Clone: CK3-6H5, Isotype: Mouse IgG1), CD13-PE (Clone: REA263, Isotype: Recombinant human IgG1), CD10-PE-Vio770 (Clone: 97C5, Isotype: Mouse IgG1), CD326-PE (Clone: HEA-125, Isotype: Mouse IgG1) and CD227-PE-Vio770 (Clone: REA448, Isotype: Recombinant human IgG1) (all Miltenyi Biotec GmbH, Bergisch Gladbach, Germany) and DAPI (Sigma-Aldrich, Hamburg, Germany).

### Microscopy

Samples were prepared using the revised staining protocol as stated above and subsequently purified using a Sony MA900 Multi-Application Cell Sorter (Sony Group Corporation, Minato, Tokio, Japan). The resulting cell suspension was sorted directly on a Polysine adhesion slide (Thermo Fisher Scientific, Waltham, USA), dried at room temperature (RT) in the dark and mounted with ProLong Diamond Antifade Mountant solution (Thermo Fisher Scientific, Waltham, USA). A Keyence BZ-9000 Biorevo fluorescence microscope (Keyence, Osaka, Japan) with DAPI and GFP filters was used for fluorescence imaging. No additional staining was carried out for the latter process. Basic image processing including cropping, brightness/contrast adjustment and gamma correction were conducted with ImageJ 1.53k.^28^

For electron microscopic analysis, cells were fixed with 2.5 % glutaraldehyde in 0.1 M sodium cacodylate buffer for 30 min at RT and stored at 4°C. Prior to EM dTEC were isolated using FACS Sorting, as CD326 was denatured by the fixation process, cells were only stained for cytokeratin, DAPI and CD227. It should be pointed out that based on our FACS data, the analyzed urine samples almost never contained CD227^+^ cell populations that stained negative for CD326. The samples were postfixed with 1 % osmium tetroxide (Electron Microscopy Sciences, Hatfield, USA) and 0.8 % potassium ferrocyanide II (Roth, Karlsruhe, Germany) in 0.1 M cacodylate buffer for 1.5 h and pelleted in agarose overnight. After dehydration in a graded ethanol series, the samples were embedded in Epon resin (Roth, Karlsruhe, Germany). Finally, ultrathin sections of the samples (70 nm) were stained with uranyl acetate, and lead citrate. The examination was carried out with a Zeiss 906 electron microscope (Carl Zeiss, Oberkochen, Germany) at 80 kV acceleration voltage equipped with a slow scan 2K CCD camera (TRS, Moorenweis, Germany).

### Statistical analysis

GraphPad Prism 9 (GraphPad Software, San Diego, California, USA) was used for calculation of median values, Spearman correlations, ROC curves, and Kaplan-Meier curves. The following statistical tests were performed: Mann-Whitney tests for the evaluation of differences in urinary cell counts of each study subgroup and Gehan-Breslow-Wilcoxon tests for the evaluation of differences in Kaplan-Meier curves for the outcome data. The significance threshold was set at p<0.05.

## Results

For detection of urinary TEC and their differentiation into cell subsets originating from the proximal or the distal kidney tubule, respectively, two antibody combinations were applied. Cytokeratin^+^CD10^+^ cell populations were regarded as proximal TEC (pTEC), while Cytokeratin^+^CD326^+^ cell populations were regarded as distal TEC (dTEC) in the exploratory study. For further improvement of differentiating urinary TEC and increasing signal-to-noise ratio of flow cytometric analysis, both marker combinations were revised for the confirmatory study. Cytokeratin^+^CD10^+^CD13^+^ cell populations were regarded as pTEC and Cytokeratin^+^CD326^+^CD227^+^ cell populations were regarded as dTEC in the confirmatory study. Additionally, DAPI was used to detect DNA-containing cells and to exclude non-DNA-containing cell debris. Analysis of urine from AKI patients showed enrichment of both cell subsets *(figure 1A+B)*. Exemplary dot-plots of control samples used to exclude background signaling can be found in *supplementary figure S1*.

**Figure 1.**
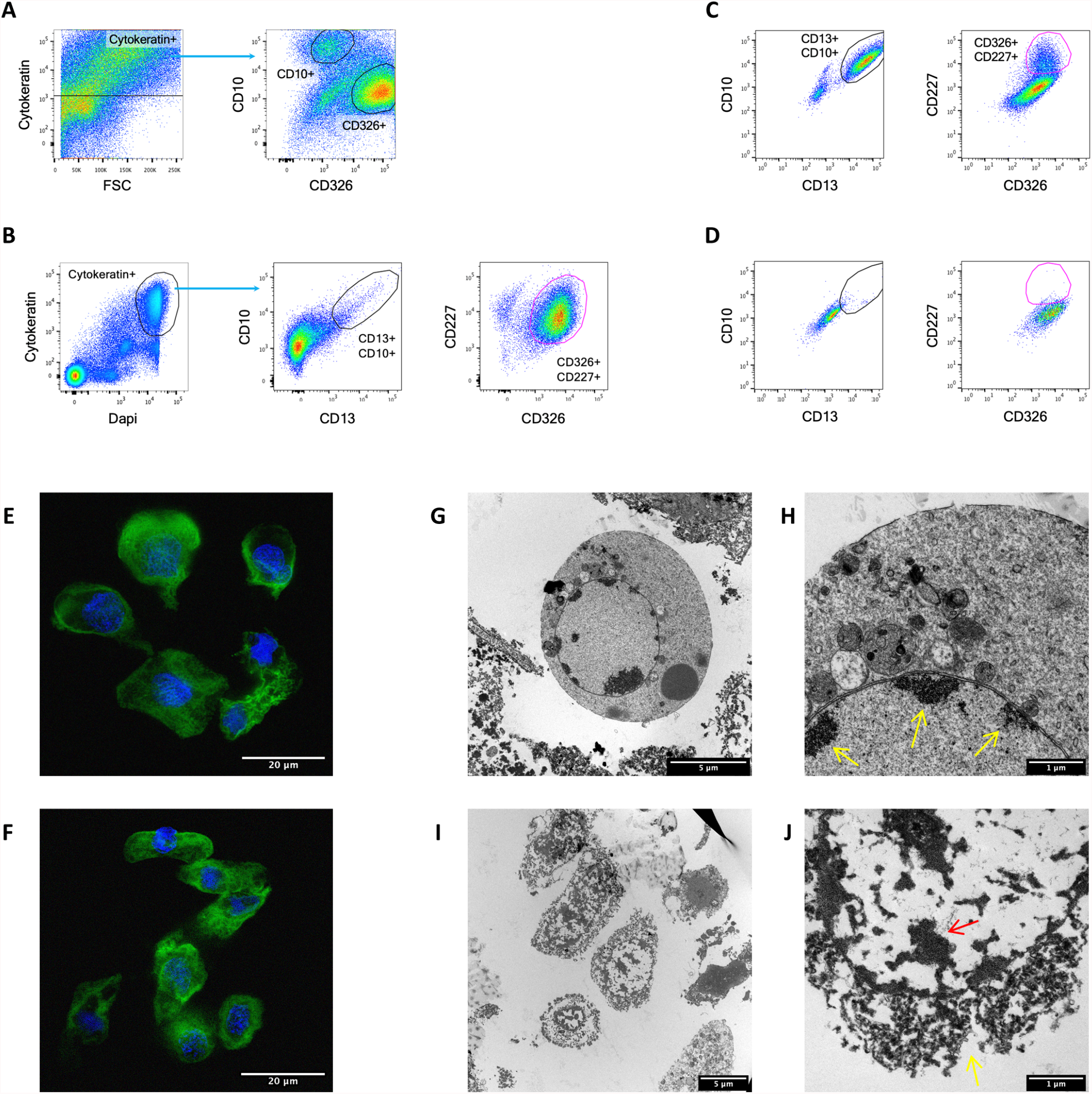
The amount of urinary tubular epithelial cells (TEC) in patients with acute kidney injury (AKI) can be measured using flow cytometry. **(A and B)** Example dot-plots for the gating strategy used in the (A) exploratory study showing proximal (CD10^+^) and distal (CD326^+^) TEC, gated on cytokeratin positive cells using unstained controls, and in the (B) confirmatory study, where proximal (CD13^+^CD10^+^) and distal (CD326^+^CD227^+^) TEC can be found, each gated on cytokeratin positive cells using FMO controls. **(C and D)** Dot-plots of a suspended post-mortem (C) renal tissue sample, showing CD13^+^CD10^+^ and CD326^+^CD227^+^ cell populations, and of a post-mortem (D) ureteral tissue sample, indicating the absence of CD13^+^CD10^+^ or CD326^+^CD227^+^ cell populations. Both gated on cytokeratin positive cells using FMO controls. **(E and F)** Fluorescence microscopic image of (E) purified Cytokeratin^+^DAPI^+^CD13^+^CD10^+^ cells and (F) purified Cytokeratin^+^DAPI^+^CD326^+^CD227^+^ cells (blue = DAPI, green = cytokeratin). **(G to J)** Electron microscopic images of purified Cytokeratin^+^CD227^+^ cells using a cell sorter. (G and detail H) Apoptotic cell with characteristic marginalization of condensed chromatin (yellow arrows in H). (I and J) Necrotic cells with chromatin clumping (red arrow in J) and loss of plasma membrane integrity (yellow arrow in J).

To control staining specificity, cells directly isolated from human tissue were analyzed. Cells isolated from post-mortem human kidney tissue (n=4) revealed staining of pTEC and dTEC for the stated dual marker combinations, whereby suspended post-mortem human ureteral tissue did only show positive signaling for CD326 (n=1). FMO controls were used to demonstrate the specificity of antibody binding *(figure 1C+D)*.

Finally, we investigated the cell morphology of the urinary TEC. Isolated Cytokeratin^+^CD13^+^CD10^+^ and Cytokeratin^+^CD227^+^CD326^+^ cells from the urine of AKI patients showed a typical tubular epithelial cell morphology on fluorescence microscopic imaging *(figure 1E+F)*. Electron microscopy of purified urinary Cytokeratin^+^CD227^+^ cells from AKI patients revealed a predominant fraction of apoptotic and necrotic cells within the sorted cell population including morphological characteristics like chromatin condensation at the nuclear periphery, rounded cell shape and apoptotic bodies as well as loss of cell membrane integrity, vacuolization and karyopyknosis. TEC in unsorted urine samples demonstrated similar features, ruling out induction of these changes during the cell isolation process *(figure 1G to J)*. Fluorescence microscopy experiments were performed twice using pooled urine samples from all together n=5 patients, electron microscopy experiments were performed twice as well using urine samples from n=1 patient each time.

### Urinary TEC reflect the severity of kidney damage and are a potential biomarker for AKI

Urinary TEC were analyzed in 84 patients with AKI, 15 inpatient controls and 10 healthy controls, divided in an exploratory and a confirmatory cohort. Patient characteristics are given in *table 1*.

As the presence of increased amounts of TEC in urine was assumed to be a damage marker for acute kidney damage, the relative quantity of TEC per 100 mL urine was correlated with the highest KDIGO stage of the 72 hours before measurement for each patient. Overall, patients with AKI showed a significantly higher count of urinary TEC compared to control groups in the exploratory study (pTEC p=0.0007, dTEC p=0.0049). This finding was confirmed in the confirmatory study (pTEC p<0.0001, dTEC p<0.0001). Patients with KDIGO stage 3 had significantly higher urinary TEC counts in comparison to patients with KDIGO stage 1 in both studies (exploratory study: pTEC p=0.0052, dTEC p=0.01; confirmatory study: pTEC p=0.0087, dTEC p=0.0026). A significant difference between the healthy control group and inpatient control group was found for proximal TEC in the exploratory cohort (pTEC: p=0.02, dTEC p=0.06), which was not observed in the confirmatory cohort (pTEC: p=0.95, dTEC p=0.79). All results are displayed in *figure 2 (A-D)*, detailed results of statistical testing can be found in *supplementary table S1*. In the confirmatory study, the amount of both urinary pTEC (p=0.01, Spearman) and dTEC (p=0.0049, Spearman) furthermore correlated with the magnitude of SCr rise (the difference between baseline SCr value and maximum SCr value of the seven days following AKI onset) (*supplementary figure S2*) and patients with septic cause of AKI (n=12; urosepsis excluded) had an overall higher quantity of pTEC (p=0.0044) and dTEC (p=0.03) compared to AKI patients with underlying cardiac condition, volume depletion or postsurgical cause (n=48). This finding can most likely be ascribed to the fact that patients with septic cause of AKI included patients with higher KDIGO stages (p=0.0002). The corresponding data is shown in *supplementary figure S3*.

**Figure 2.**
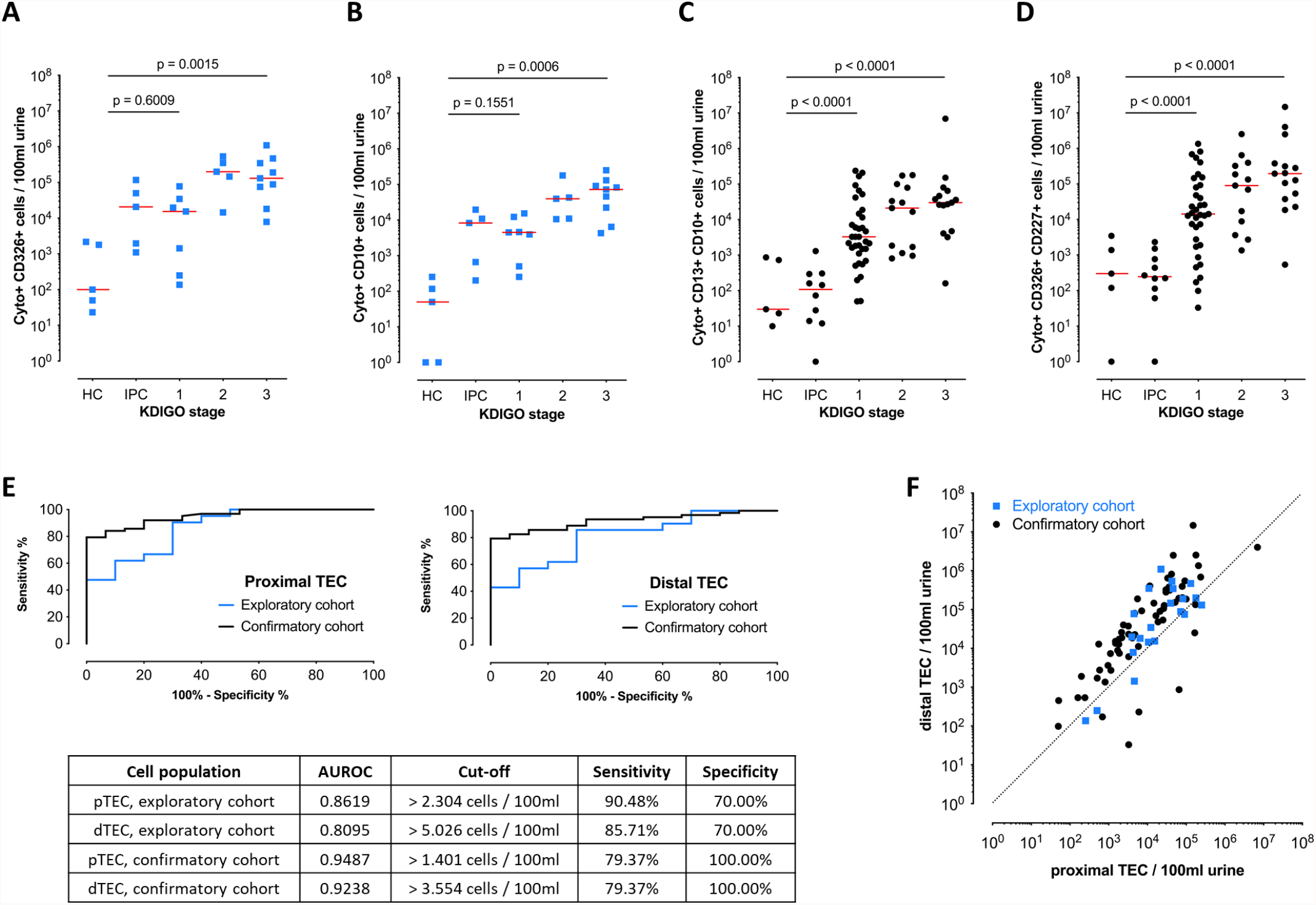
Using flow cytometry, the amount of urinary TEC functions as a biomarker for the detection of acute kidney injury (AKI). HC, healthy control; IPC, inpatient control; 1-3, KDIGO stage of patients with AKI. Control groups were pooled for comparison calculations. **(A and B)** Amount of proximal TEC (Cytokeratin^+^CD10^+^) and distal TEC (Cytokeratin^+^CD326^+^) in the exploratory cohort with n=21 patient samples, n=5 healthy control samples and n=5 control samples from inpatients without AKI. **(C and D)** Amount of proximal TEC (Cytokeratin^+^CD13^+^CD10^+^) and distal TEC (Cytokeratin^+^CD326^+^CD227^+^) in the confirmatory cohort with n=63 patient samples, n=5 healthy control samples and n=10 inpatient control samples. Comparisons calculated by Mann-Whitney test, for detailed results of statistical testing see *Supplementary table S1*. **(E)** Receiver-operator characteristic (ROC) analysis for urinary proximal and distal TEC comparing patients with AKI and patients of both control groups used in the exploratory (blue) and confirmatory (black) study. Area under the curve (AUROC) for proximal TEC: 0.8619 (exploratory study); 0.9487 (confirmatory study). AUROC for distal TEC: 0.8095 (exploratory study); 0.9238 (confirmatory study). **(F)** Correlation of urinary proximal distal TEC counts per 100 mL urine each for exploratory (blue) and confirmatory (black) cohort. The cell counts were correlated with each other with relatively higher cell counts for distal TEC. (Exploratory cohort: Spearman r=0.6935, p=0.0005; confirmatory cohort: Spearman r=0.8262, p<0.0001).

Receiver operator characteristics (ROC) curves for both urinary proximal and distal TEC were used to analyze the sensitivity and specificity for distinguishing individuals with AKI from individuals without acute kidney impairment. The relative amount of urinary pTEC and dTEC as a structural damage biomarker performed well in identifying patients with AKI regardless of the stage in the exploratory cohort, (AUC=0.8619 for pTEC, AUC=0.8095 for dTEC) while the results were even better in the confirmatory cohort (AUC=0.9487 for pTEC, AUC=0.9238 for dTEC). Cut-off values calculated with Youden’s test as well as all ROC curves are displayed in *figure 2E*.

Comparing the collected data for proximal and distal TEC counts of each patient showed a correlation between urinary proximal and distal TEC amounts, with a predominant distal TEC fraction. Spearman test revealed a strong correlation for the exploratory study data (r=0.6935, p=0.0005, median of dTEC/pTEC quotients = 2.3) and an even stronger correlation for the confirmatory study data (r=0.8262, p<0.0001, median of dTEC/pTEC quotients = 5.0) (*figure 2F*). Further analysis of patients with predominant proximal TEC counts did at this point not provide a general difference in patient’s gender, age, or etiology of AKI.

### Urinary TEC predict the outcome of AKI

Compared to healthy and inpatient controls, a fraction of AKI patients did not present elevated amounts of TEC in their urine (confirmatory cohort data; fraction of AKI patients with TEC counts below maximum TEC count of healthy cohort: 16% for pTEC, 21% for dTEC). These patients showed a quicker AKI recovery than patients with increased amounts of urinary TEC in the confirmatory cohort (pTEC cut-off 863/100ml, p=0.02; dTEC cut-off 3472/100ml urine, p=0.03; cut-off values determined according to the cell counts in healthy controls). However, this was not observed when only patients with KDIGO stage 1 were included in the analysis (pTEC p=0.23, dTEC p=0.30). Shorter recovery timespans could thus be due to less severe kidney injury implied by lower KDIGO stage (*figure 3A-D*). This analysis was omitted in the exploratory cohort owed to the low number of AKI-patients with normal range TEC (n=1 for pTEC, n=3 for dTEC).

**Figure 3.**
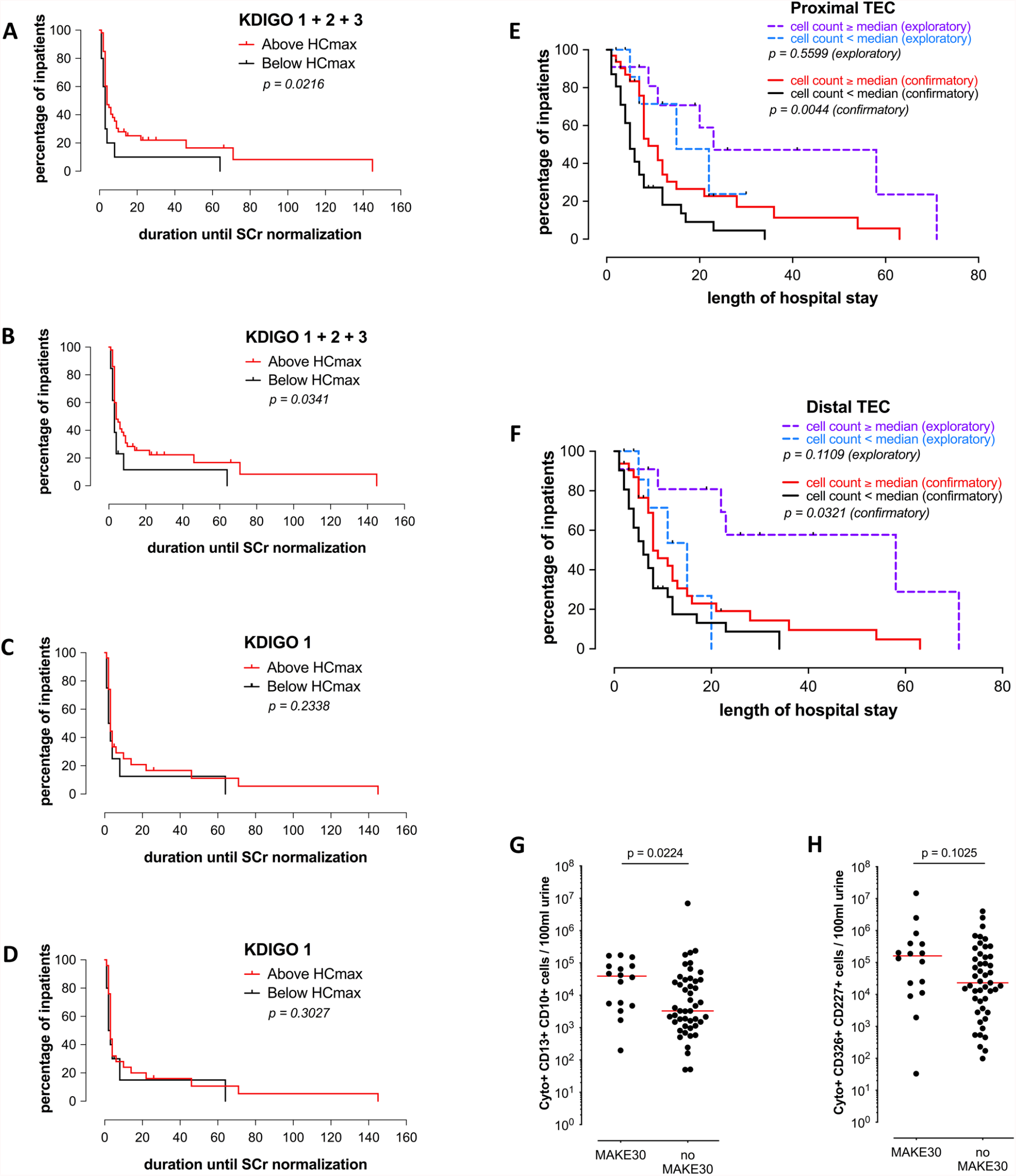
Urinary TEC as a biomarker for the outcome of acute kidney injury. **(A to D)** Kaplan-Meier curves of confirmatory cohort describing time to recovery defined by normalization of SCr. AKI patients of all stages with cell counts above the maximum cell count of healthy controls (red) were less likely to recover quickly than patients with TEC counts comparable to healthy controls (black). (**A** proximal TEC *p=0.02, **B** distal TEC *p=0.03). The difference was not significant if only patients with AKI stage 1 were included (**C** proximal TEC p=0.23, **D** distal TEC p=0.3). **(E and F)** Kaplan-Meier curves describing the length of hospital stay from the day of AKI onset (based on the KDIGO definition of AKI) in dependence of the urinary TEC count. Tick marks indicate death of patients. Patients with higher cell counts were more likely to stay longer in hospital until they got dismissed or died (proximal TEC: Exploratory cohort: p=0.56, Confirmatory cohort: **p=0.0044; distal TEC: Exploratory cohort: p=0.11, Confirmatory cohort: *p=0.03; Gehan-Breslow-Wilcoxon-test). **(G and H)** Correlation of urinary TEC counts and the appearance of MAKE30 events in the confirmatory study. Patients with a MAKE30 event after AKI onset had relatively higher proximal TEC counts compared to patients without MAKE30 event (proximal TEC: *p=0.02; distal TEC: p=0.10; Mann-Whitney test)

As a second outcome parameter we tested whether urinary TEC counts are able to predict the timespan from urinary TEC measurement to hospital discharge as proxy for disease severity. Patients with cell counts above the median stayed significantly longer in hospital after urine measurement than patients with cell counts below the median. This outcome was observed for both pTEC (p=0.0044) and dTEC (p=0.03) in the confirmatory study but not in the exploratory study (pTEC: p=0.56; dTEC: p=0.11), which might partly be due to the small cohort size of this first study (*figure 3E+F*). The significant difference was also detectable when only patients with KDIGO 2 and 3 were included for Gehan-Breslow-Wilcoxon testing (pTEC and dTEC p=0.0360).

Lastly, urinary TEC counts were tested for prediction of the well-established MAKE-30 outcome criteria. The ratio of MAKE-30 occurrence to non-occurrence was 25%:75% in the confirmatory study. Patients of the MAKE-30 group (n=16) had significantly higher urinary proximal TEC counts (p=0.02) but not significantly higher urinary distal TEC counts (p=0.10) than the patients of the non-MAKE-30 group (n=47) (*figure 3G+H*). Area under the ROC curve for occurrence of MAKE-30 was 0.69 for pTEC, 0.63 for dTEC, which was superior to SCr (AUC 0.62) and eGFR (AUC 0.60) on the day of measurement.

### Amount of urinary TEC decreases over time after AKI

To assess the changes in urinary TEC counts over time after AKI, we performed follow-up measurements for a small sub-cohort of 7 patients (exploratory cohort n=4, confirmatory cohort n=3). Overall, cell counts for pTEC as well as for dTEC decreased after AKI onset (*figure 4*). In addition, urine samples from patients that had already recovered from AKI within 48 hours before measurement showed lower TEC counts. Despite recovery, some patients retained elevated amounts of urinary TEC for several days.

**Figure 4.**
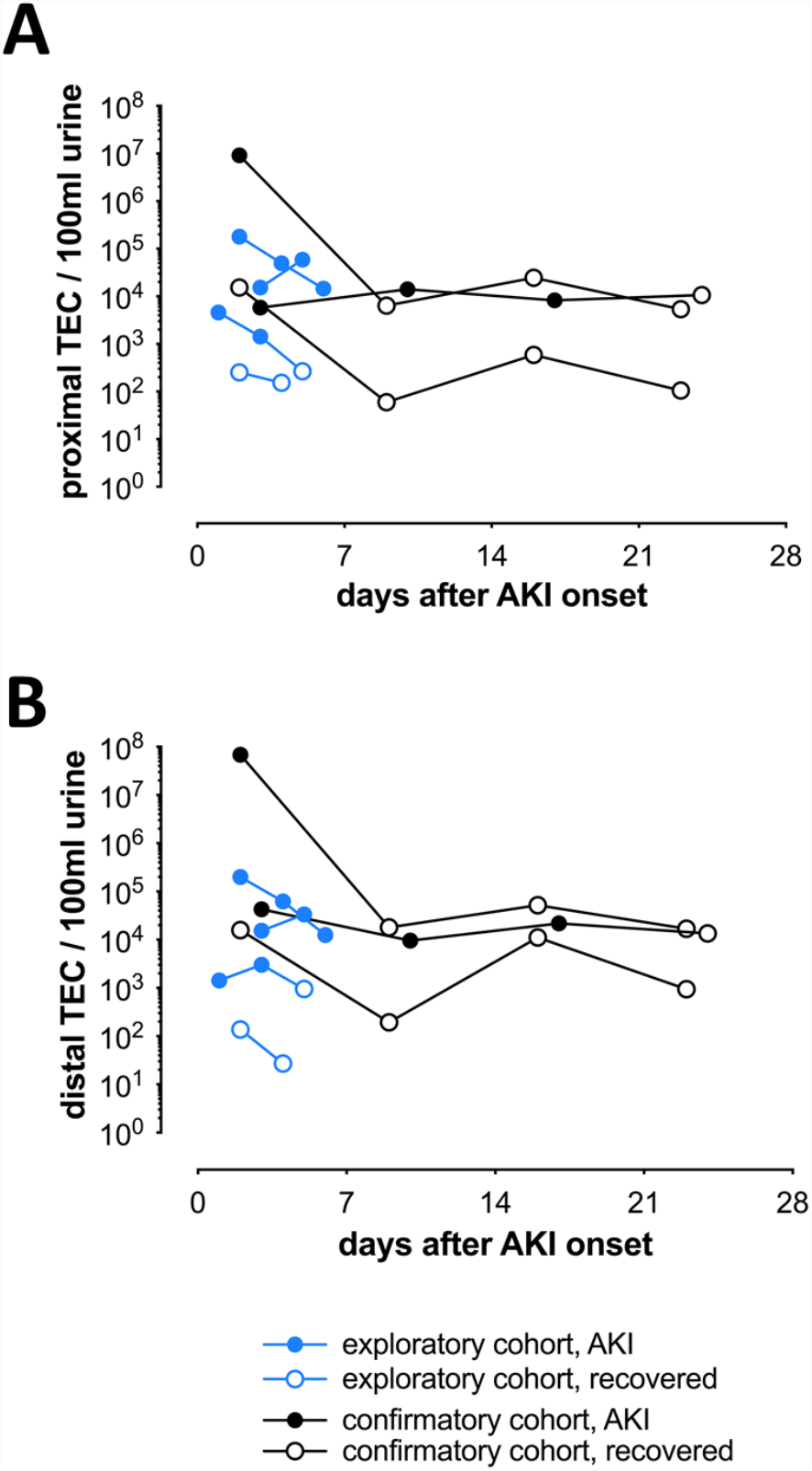
Follow-up controls of urinary TEC counts of patients with AKI from the exploratory (blue, n=4) and confirmatory (black, n=3) cohort, plotted against time after AKI onset. Full circles indicate fulfillment of KDIGO criteria for AKI based on the initial baseline serum creatinine, empty circles indicate recovery to normal serum creatinine values. **(A and B)** Proximal and distal TEC counts assumably tend towards an individual normalized level.

## Discussion

In this paper we introduce the flow cytometric detection of urinary TEC for monitoring kidney damage in patients with AKI. Moreover, we demonstrate the correlation between their relative quantity and AKI severity. Even though earlier microscopic studies have already found correlations between urinary TEC enrichment and severity of AKI,^16–18^ this is the first report using a flow cytometric approach to assess the extent of structural damage in the acutely injured kidney. Previous work from our group introduced the possibility of detecting tubular cells in urine via flow cytometry and found the combination of TEC and T cell counts to be predictable for acute kidney transplant rejection.^20^ We conducted two consecutive cross-sectional studies to investigate the initial TEC marker combination as well as a revised version for their ability of monitoring tubular damage and outcome during AKI. Cytokeratin, CD10 and CD326 proved to be feasible for detecting proximal and distal tubular cells and their amounts reached significantly increasing values with higher underlying KDIGO stages in the exploratory cohort, the addition of CD13 and CD227 refined the staining in the larger confirmatory cohort by improving the distinction of TEC from other urinary parts and background noise, which is a major limitation in flow cytometric analyses.

Patients with increased urinary TEC counts were characterized by higher KDIGO stages, SCr magnitude, later recovery, prolongated hospital stay, and more frequent MAKE-30 occurrence. The amount of urinary pTEC was of similar predictive value for MAKE-30 events as urinary NGAL.^29^ The observation of a predominance of dTEC within urine samples joins the debate about the main site of tubular damage in the context of AKI. Assuming that proximal tubules are more sensitive to cell death, especially after ischemia, these findings are contrasting.^30^ A possible explanation for this paradox might be a better preservation of surface markers on dTEC and the shorter distance that shedded dTEC need to “travel” before detection in the voided urine. In addition, the proportion of the tubular and collecting duct system expressing CD326 and CD227 is overall higher than the one expressing CD13 and CD10.^25,31^ Proximal TEC yet predicted outcome more significantly. The informative value of the respective pTEC:dTEC ratios lastly remains unclear. Our longitudinal assessment only allows cautious conclusions to be drawn about the diagnostic value of urinary TEC in the recovery phase after acute kidney injury. Nonetheless, they suggest a normalization over the course of a few weeks, which is in accordance with the damage-repair model of tubular injury.^30^

Urinary TEC counts most likely reflect the extent of tubular death,^32^ therefore we propose urinary TEC as possible biomarkers to distinguish prerenal azotemia (PRA) from acute tubular injury (ATI), two conditions that currently rely on creatinine increase, urine output and sediment microscopy, but have potential clinical implications.^33^AKI patients without elevated amounts of urinary TEC might represent the classical PRA patients, with absence of structural damage and quick recovery. However, there was no measurable difference in damage severity or recovery of AKI stage 1 patients with or without elevated TEC. Furthermore, the observed continuous increase of urinary TEC with progressing AKI stages challenges the PRA-ATI dichotomy but suggests a rather continuous damage spectrum in AKI.

Flow cytometric urinalysis finally contains the opportunity of using the identified TEC as a platform for individual characterization of different types of tubular dysfunction based on the cellular level. Further phenotyping of urinary TEC by complementing the established staining protocol with additional markers for activation, proliferation, senescence or cell death would provide valuable insights for monitoring kidney function.

Limitations of the studies presented here include small cohort size and the single-center cross-sectional design with the restriction to SCr values measured in routine clinical practice. By assessing our biomarker in relation to serum creatinine, its validity is restricted to the bias that is inherent in creatinine use. Variables like urine dilution, comorbidities, intake of nephrotoxic drugs, different AKI etiologies and initiation of volume therapy should be taken into consideration in further investigations and a more comprehensive kinetic characterization is to be aimed for. Despite all these constraints, flow cytometric urinalysis of TEC holds the ability to quantify current extent of tubular damage in a non-invasive, observer-independent fashion.

## Data Availability

All data produced in the present study are available upon reasonable request to the authors.

## Acknowledgements

We would like to thank the Flow Cytometry & Cell Sorting Facility (FCCF) of the DRFZ for technical support and helpful insights.

This work was supported by a grant from the Jackstädt-Stiftung and funds from BIH Berlin.

JK was supported by a research scholarship of the Deutsche Gesellschaft für Nephrologie (DGfN). VL was supported by a research scholarship of the SFB650 at Charité-Universitätsmedizin Berlin.

## Supplementary Material

**Supplementary figure S1** Exemplary dot-plots for distinction between positive and background signals in flow cytometric detection of tubular epithelial cells using control samples. (PDF)

**Supplementary figure S2** Correlation of TEC counts per 100 mL urine and SCr magnitude (PDF)

**Supplementary figure S3** Correlation of proximal and distal TEC counts with etiology of AKI. (PDF)

**Supplementary table S1** Detailed results of statistical testing from correlation of the amount of urinary TEC and AKI severity. (PDF)

