## Supplemental Material for "Monitoring tubular epithelial cell damage in AKI via urine flow cytometry"

*SUPPLEMENTARY FIGURE S1*

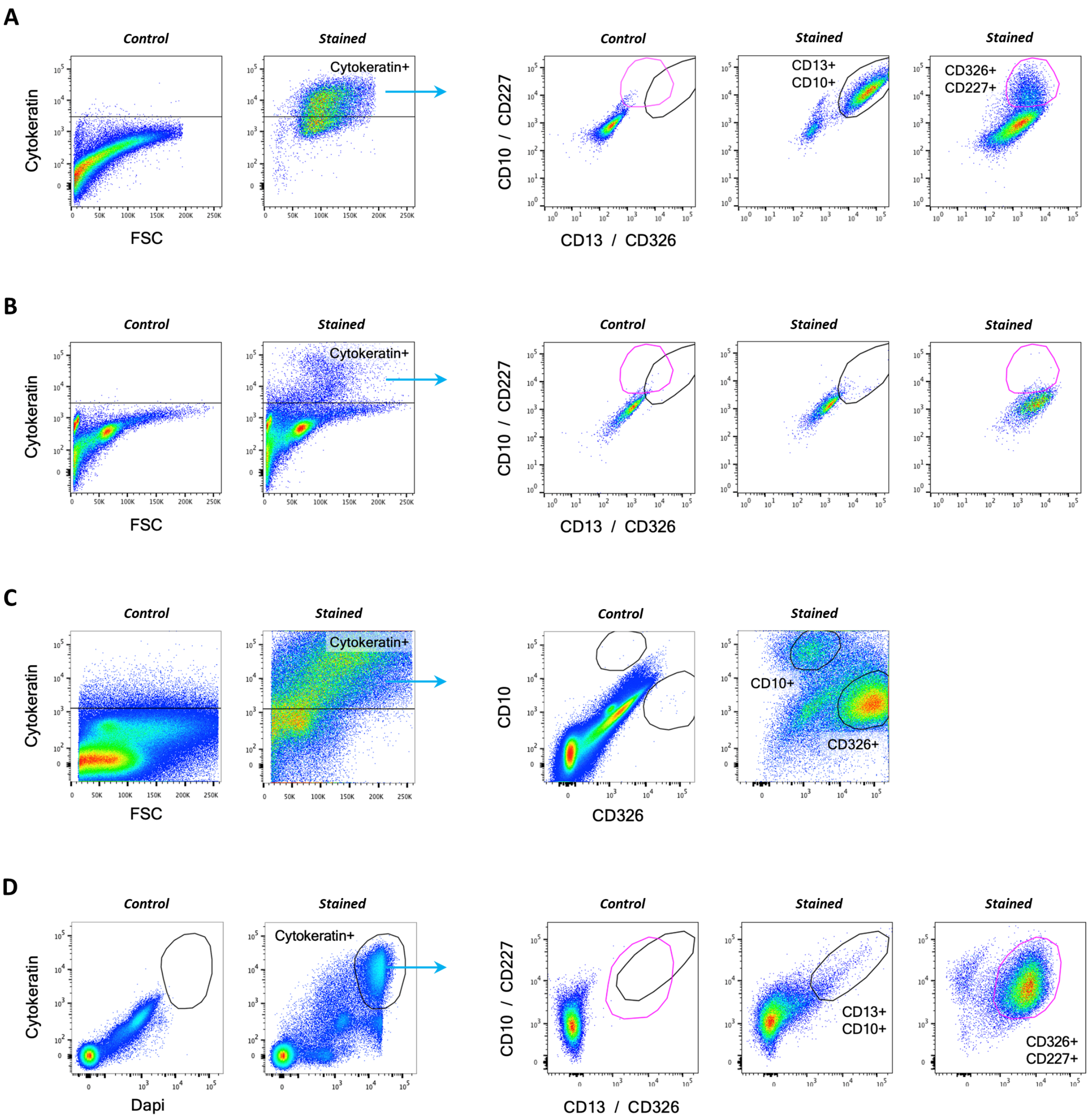


**Supplementary Figure S1** Exemplary dot-plots for distinction between positive and background signals in flow cytometric detection of tubular epithelial cells using control samples. **(A and B)** Dot-plots of (A) suspended kidney tissue sample and (B) suspended ureteral tissue sample. FMO controls containing cytokeratin and isotype antibodies were used. **(C)** Dot-plots of a sample from the exploratory cohort. Unstained controls were used to determine background signaling. **(D)** Dot-plots of a sample from the confirmatory cohort. FMO controls containing cytokeratin-APC and isotype antibodies for CD10-PE-Vio770 and CD13-PE or CD227-PE-Vio770 and CD326-PE were used to determine background signaling.

*SUPPLEMENTARY FIGURE S2*

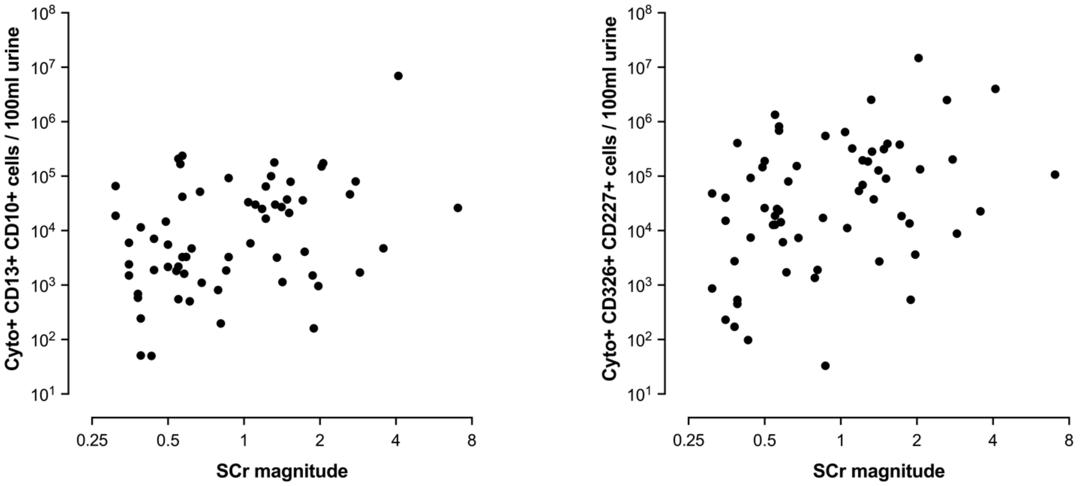


**Supplementary Figure S2** Correlation of TEC counts per 100 mL urine and SCr magnitude (difference between baseline SCr to maximum SCr of the 7 days after AKI onset). Both proximal (Spearman r=0.3180, *p=0.01) and distal (Spearman r=0.3500, **p=0.0049) TEC counts correlate with the SCr magnitude. Data from confirmatory cohort.

*SUPPLEMENTARY FIGURE S3*

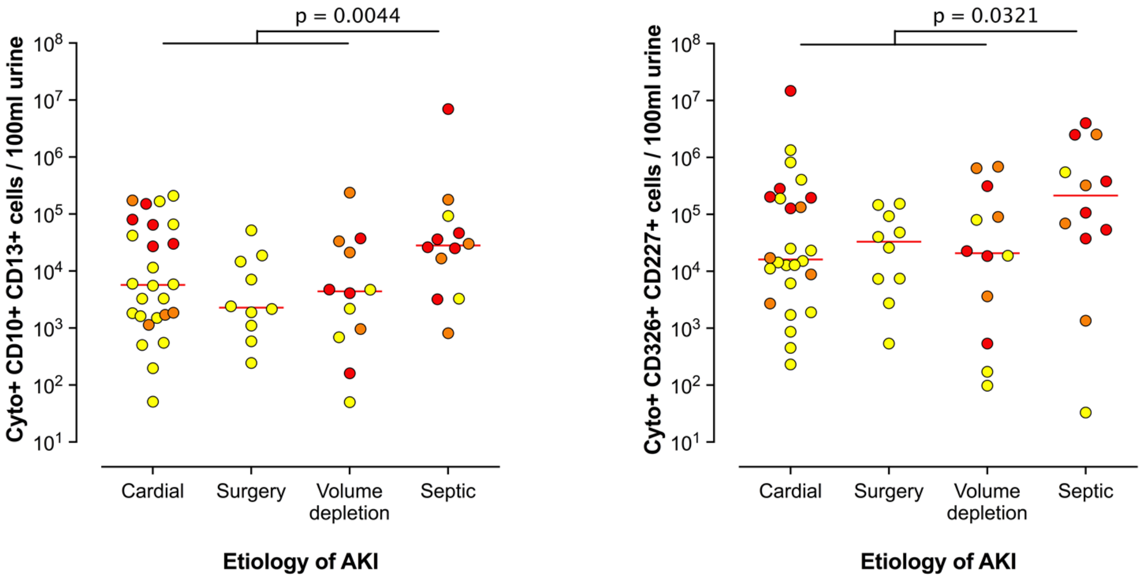


**Supplementary Figure S3** Correlation of proximal and distal TEC counts with etiology of AKI. Patients with sepsis as underlying etiology of AKI had significantly higher proximal (**p=0.0044) and distal (*p=0.03) TEC counts than patients with cardiac cause, post-surgery patients or patients with AKI due to volume depletion (Mann-Whitney-test). Colors indicate KDIGO stage (yellow = KDIGO 1, orange = KDIGO 2, red = KDIGO 3).

*SUPPLEMENTARY TABLE S1*

| **A** | Controls – KDIGO 1 | Controls – KDIGO 2 | Controls – KDIGO 3 | KDIGO 1 – KDIGO 2 | KDIGO 1 – KDIGO 3 | KDIGO 2 – KDIGO 3 |
|---|---|---|---|---|---|---|
| pTEC, exploratory cohort | 0.1551 | 0.0047 | 0.0006 | 0.3030 | 0.0052 | 0.6064 |
| dTEC, exploratory cohort | 0.6009 | 0.0047 | 0.0015 | 0.0303 | 0.0115 | 0.6064 |
| pTEC, confirmatory cohort | < 0.0001 | < 0.0001 | < 0.0001 | 0.1520 | 0.0087 | 0.4956 |
| dTEC, confirmatory cohort | < 0.0001 | < 0.0001 | < 0.0001 | 0.1390 | 0.0026 | 0.3627 |

<table>
<tr><th colspan="3">B</th><th>Healthy controls (HC)</th><th>Inpatient controls (IPC)</th><th>KDIGO 1</th><th>KDIGO 2</th><th>KDIGO 3</th></tr>
<tr><td rowspan="4">Exploratory cohort</td><td rowspan="2">Median</td><td>proximal TEC</td><td>50</td><td>8.267</td><td>4.500</td><td>39.532</td><td>72.331</td></tr>
<tr><td>distal TEC</td><td>100</td><td>20.667</td><td>15.267</td><td>200.000</td><td>131.200</td></tr>
<tr><td rowspan="2">Standard deviation</td><td>proximal TEC</td><td>106</td><td>8.045</td><td>5.769</td><td>70.156</td><td>76.929</td></tr>
<tr><td>distal TEC</td><td>1.062</td><td>48.543</td><td>27.941</td><td>200.383</td><td>347.334</td></tr>
<tr><td rowspan="4">Confirmatory cohort</td><td rowspan="2">Median</td><td>proximal TEC</td><td>30</td><td>109</td><td>3.273</td><td>21.201</td><td>29.976</td></tr>
<tr><td>distal TEC</td><td>301</td><td>246</td><td>14.309</td><td>89.961</td><td>195.863</td></tr>
<tr><td rowspan="2">Standard deviation</td><td>proximal TEC</td><td>427</td><td>393</td><td>59.122</td><td>64.277</td><td>1.778.813</td></tr>
<tr><td>distal TEC</td><td>1.458</td><td>754</td><td>290.766</td><td>685.222</td><td>3.829.688</td></tr>
</table>

**Supplementary Table S1** Detailed results of statistical testing from correlation of the amount of urinary TEC and AKI severity.
**(A)** P-values of Mann-Whitney-Tests comparing TEC quantities of subgroups from the exploratory and confirmatory cohort.
**(B)** Median and standard deviation values of subgroups from the exploratory and confirmatory cohort.
